# STRENGTHENING MATERNAL REFERRAL SYSTEMS: A POLICY QUALITATIVE ANALYSIS IN INDONESIA

**DOI:** 10.1101/2025.03.27.25324569

**Authors:** Siti Rani Angelina, Likke Prawidya Putri, Laksono Trisnantoro

**Author notes:** **Corresponding Author:** Likke Prawidya Putri; Department of Health Policy and Management, Faculty of Medicine, Public Health, and Nursing, Universitas Gadjah Mada, Yogyakarta, Indonesia.

## Abstract

**Background:** Indonesia continues to face high maternal mortality rates despite efforts to strengthen maternal healthcare systems. The introduction of various referral systems, from both national and sub-national governments, were aiming to improve referral efficiency, including for maternal emergency cases. This study examines the barriers and opportunities utilizing the various referral information systems from the policy content and real-world implementation.

**Methods:** We applied a qualitative policy analysis by analyzing the content of the existing policies and interviewing key informants working in community health centers, hospitals, and district authorities. Content analysis was performed towards the policy documents and the interview transcripts.

**Results:** The key barriers in implementing maternal referral systems include standard and monitoring, quality of care, and communication. There were lacking standard on definition of cases for referral as well as classification of health facilities capable to treat maternal and neonatal emergency cases. The monitoring of systems utilization was limited. The standardized emergency maternal training should be provided for both health and non-health staff involved in the maternal referral systems.

**Conclusion:** To enhance usefulness and effectiveness of maternal referral systems, health authorities should classify maternal cases by severity, map facility capacities, establish clear communication guidelines, and provide integrated training covering clinical, managerial, and digital skills. Regular monitoring should be conducted to refine program implementation and improve maternal health outcomes.

## INTRODUCTION

Reducing maternal mortality rate (MMR) remains a longstanding challenge in Indonesia. While the country successfully lowered the MMR from 299 per 100,000 live births in 2000 to 159 in 2019, the rate increased again to 173 in 2020. The rate was also comparably higher than other Southeast Asian countries with similar income levels, such as Vietnam and the Philippines with MMR of 46 and 78, respectively (1). Also, the Indonesian figure is comparably high than the targeted sustainable development goals (SDGs) of 70 per 100,000 (2).

Studies indicate that the high maternal mortality rate in Indonesia stems from multiple factors, including both maternal characteristics and health system limitations. Maternal factors such as educational status and socioeconomic conditions were associated with maternal mortality (3, 4). On the healthcare system side, key determinants include the availability of doctors and midwives, geographic accessibility of the health facilities, lacking standard operational procedures, and poor quality of care (5–7). Additionally, the delays to seek care has been identified as a fundamental contributor to the persistently high maternal mortality rate (8–10).

Delays leading to complications could be prevented through timely treatment and high-quality care. One of the key strategies that has been implemented to reduce referral delays in Indonesia is the introduction of the Maternal and Neonatal Referral Manual (11). A maternal referral system allows for identifying and classifying risks of maternal and neonatal emergency cases, then plan and design the treatment for such emergency cases strategically, proactively, and pragmatically to ensure that all maternal emergency cases receive prompt and effective treatment. Previous studies have shown that maternal mortality rates are higher among cases with delayed referrals compared to those with timely referrals (12, 13). The implementation of the referral manual aligns with the government’s efforts to expand and strengthen Basic Emergency Obstetric and Neonatal Care (BEONC) and Comprehensive Emergency Obstetric and Neonatal Care (CEONC) facilities. Several initiatives have integrated the maternal referral manual into the routine and emergency referral procedures, incorporating elements such as the use of text messaging or mobile applications for maternal emergency case reporting and the development of standardized maternal emergency management guidelines in primary healthcare services (14, 15).

Since the introduction of maternal referral systems in Indonesia in the 2010s, the government has implemented several initiatives aimed at enhancing the referral process for general medical conditions, that may include maternal and neonatal cases, including the *Sistem Informasi Rujukan Terintegrasi* (SISRUTE) and the *Sistem Penanggulangan Gawat Darurat Terpadu* (SPGDT). SISRUTE, launched in 2016, is an online-based referral information system that maps healthcare facilities and provides recommended patient referral pathways from primary care to referral care facilities, thus streamlining the referral process (16). SPGDT, on the other hand, is an integrated emergency response system designed to provide rapid and effective healthcare interventions in any emergency situations (17). While both systems map healthcare facilities for emergency case management, SPGDT differs from SISRUTE in that it is accessible to the general public, whereas SISRUTE is exclusively available to healthcare facilities managing patient referrals. SPGDT is also integrated with the national 119 emergency hotline, which receives emergency reports from patients (18, 19).

In addition to the national government’s introduction of SISRUTE and SPGDT, several subnational governments have developed their own referral systems as part of innovative efforts to enhance healthcare services. Bogor district, which has one of the highest number of maternal mortality in Indonesia, introduced SITEGAR, a systems that facilitates patient referrals from primary care to hospitals. SITEGAR facilitates patient referrals from primary care to specialized care, similar to SISRUTE, but with more detailed information regarding the availability of healthcare personnel and equipment at local health facilities.

A scoping review mapped the challenges of utilizing information systems in maternal referral that encompasses patient and health systems factors (20). Align with the review’s findings, studies in Indonesia identified that family support, mother’s and family’s knowledge, and administrative procedures has been among the key barriers of mothers to be referred to the appropriate healthcare facilities (4, 21, 22). It is also important to decide on the indicators for patient condition that requires referral to improve communication between health professional in the sender and recipient facilities (23). The lack of complete information on the availability and capacity of referral health facilities was one of the obstacle of referring from primary to referral care (22).

Exploring the intricacies of maternal referral systems is the key to developing better and more effective maternal referral systems in Indonesia. It is important to further understand how the health system can be strengthened to achieve even cooler maternal referral systems. Moreover, there are various systems that have similarities and opportunities for overlap with one another. Investigating the barriers and facilitators within maternal referral systems would allow us to identify critical areas for enhancing the quality of care and reduce maternal mortality rates in Indonesia. This study aimed at analyzing the policy on the existing maternal referral systems, focusing on the context, content, and actors. This article also aims at exploring the real-world challenge of having multiple healthcare referral systems by conducting a case study in Bogor district.

## METHODS

This study employed qualitative document analysis followed by key informant interviews and observation to validate the findings from document analysis. Our analysis is focused on Bogor district as an example of a district with long-standing problem of high number of maternal mortality and multiple healthcare referral systems.

### Policy Analysis

We searched government websites for relevant policies and/or program related to maternal referral systems with the keywords ‘maternal’ AND ‘referral’ OR ‘emergency’. The manual and systems for maternal referral care was also obtained from direct preliminary interviews with the local stakeholders. Policy analysis was conducted using the policy triangle (24) combined with the framework of health systems barriers in maternal healthcare systems (20). For each identified policy / program, we further identified the content regulation and/or technical guidance to collect information on: 1) standard and monitoring, 2) communication, 3) quality of care, 4) transportation, 5) referral documentation, and 6) network infrastructures.

### Key informant interviews

The key informant interview involved stakeholders and staff that were in charge to implement the referral systems in Bogor regency, West Java, Indonesia. We purposively selected informants using the maximum variation approach according to their involvement in the maternal referral systems in the district: 2 public community health centers (Puskesmas), maternal referral hospitals, and staff of Bogor district health authority. The Puskesmas was selected according to the highest and lowest frequency of using SITEGAR in the last 6 months.

The semi-structured interview guide was developed based on the framework on barriers in maternal and neonatal referral systems (20). The interviews were conducted face-to-face, with the duration of 45 to 90 minutes each. The interviewer was a midwife, studying for Master of Health Policy and Management, residing in Bogor city, had never worked in public sector.

The interview was conducted between 13-03-2023 and 30-06-2023. Written consent was sought from all informants, with each paper-based consent form signed by the informant and one witness. All informants were over 18-year-old; hence no consent was required from parents or guardians. All informants were free to withdraw from the study at any time they wished without any change in their health care process. We assured the informants’ confidentiality. Ethical approval was obtained from the Ethics Committee for Medical and Health Research Ethics Committee (MHREC) Faculty of Medicine, Public Health and Nursing Universitas Gadjah Mada - Dr. Sardjito General Hospital (approval number: KE/FK/0364/EC/2023).

The interviews were recorded, transcribed verbatim and coded using NVivo v.14.23. Interview summaries were sent to the respondents for member checking. We employed both deductive and inductive approaches in coding the data. The content analysis was applied, using the framework of barriers to the use of information systems in maternal and neonatal case proposed by Harahap et al (2019) as the key themes and sub-themes (20). The emerged codes and sub-themes and the themes were then defined, considered, and retitled through in-depth discussion with three authors. Member checking was completed by sending the summary of interviews to the respondents.

The interviews were followed by direct observation took place for 7 days in the district hospital and each Puskesmas, and 2 weeks of observation in both Puskesmas. The checklist focused cross-checking informants’ information about the communication, standard and monitoring, referral documentation, and transportation (25).

## RESULTS

The document search identified 4 policies related to maternal referral systems that were applicable in Bogor district: SISRUTE, SPGDT, SITEGAR, and the maternal referral itself. The information about each policy is shown in Table 1.

**Table 1.**
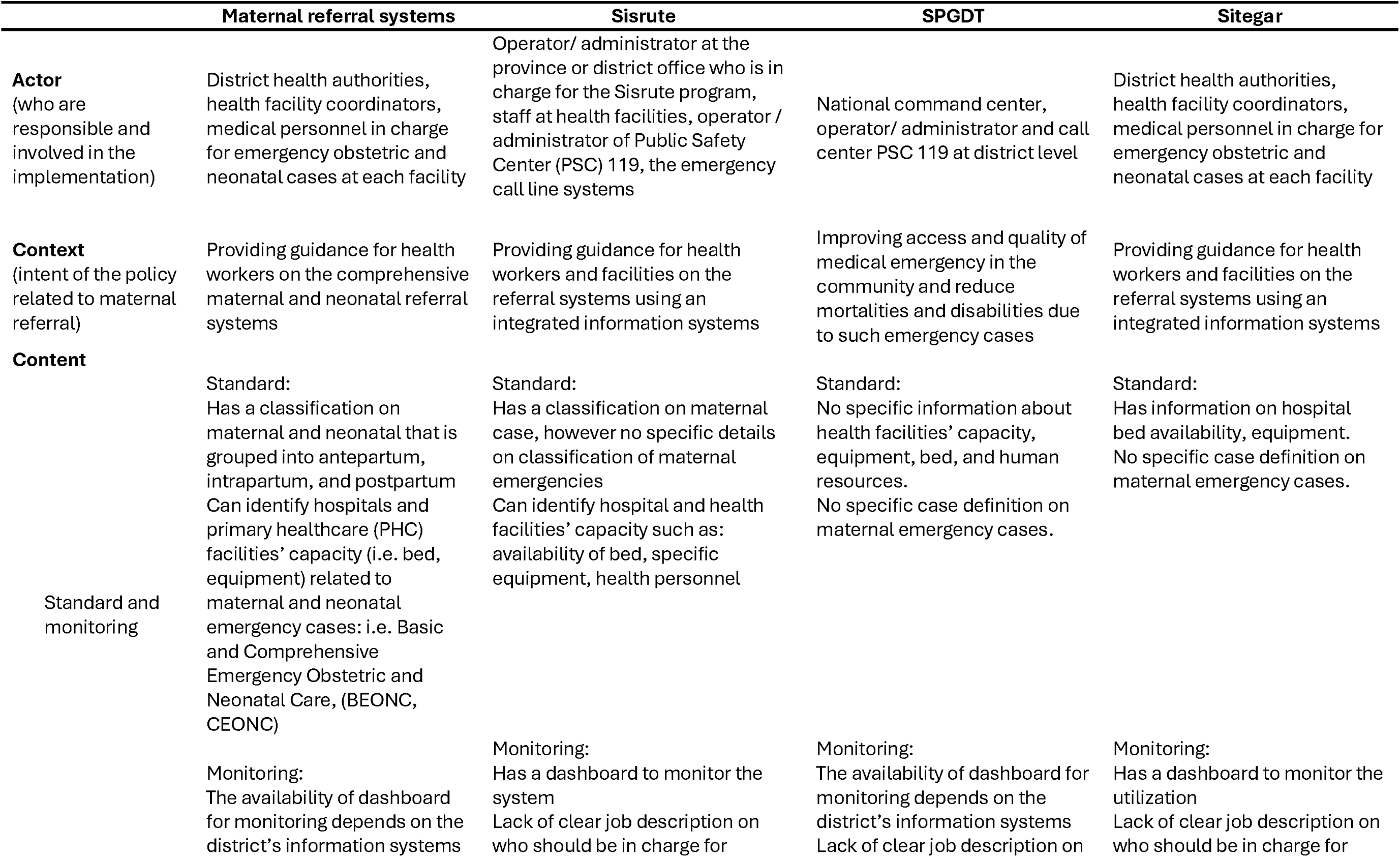
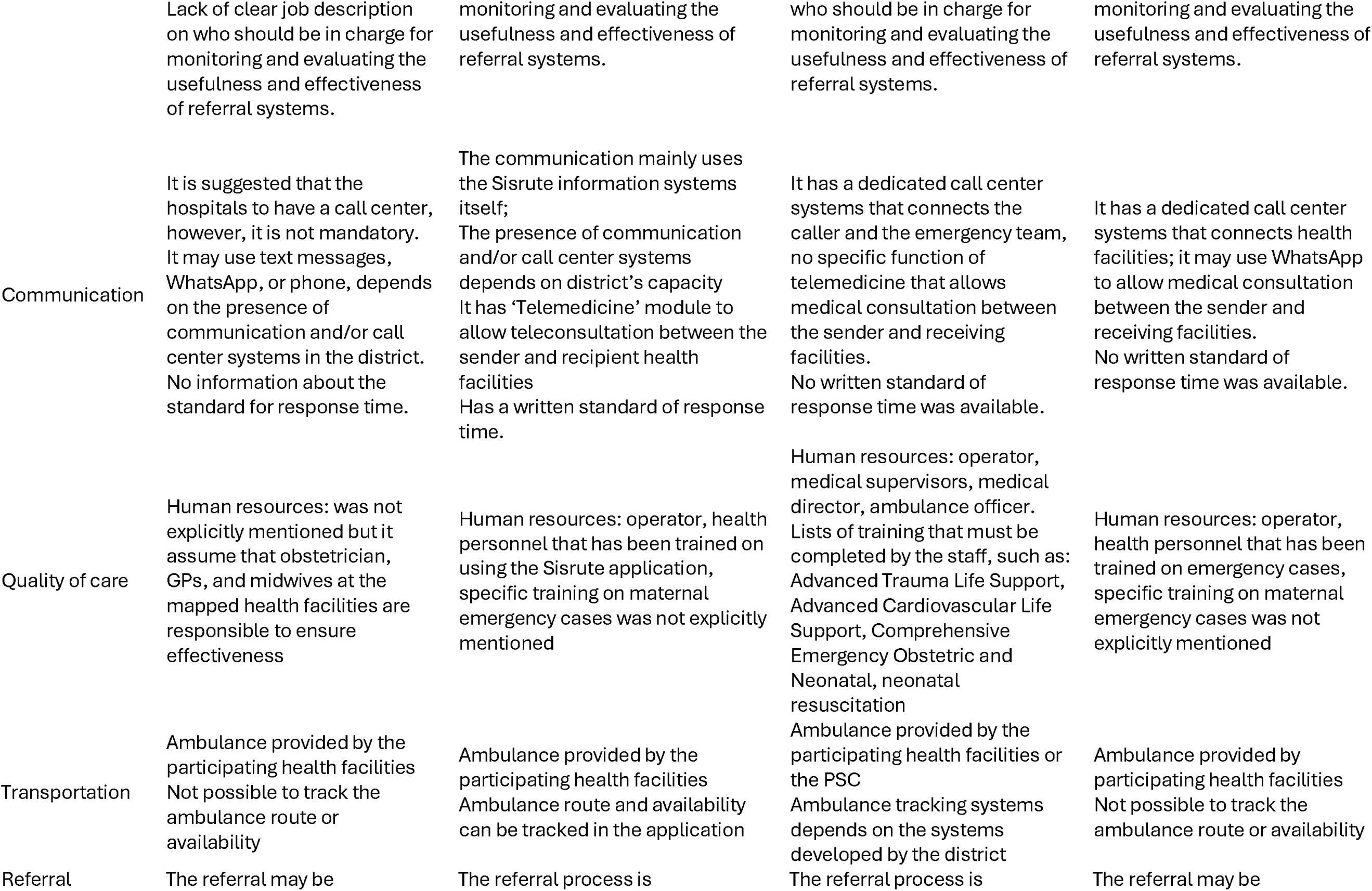
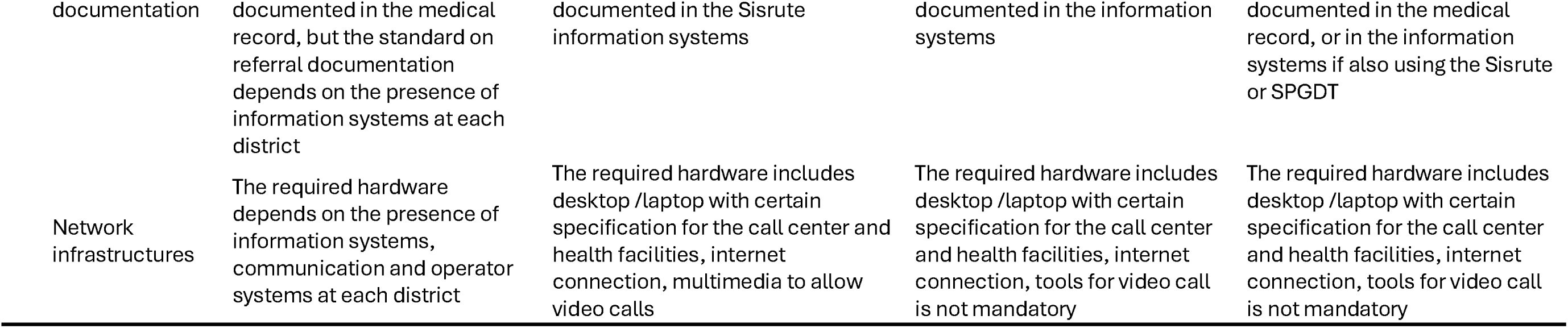
Actors, contexts, and contents of the policies related to maternal referral systems.

A total of 8 informants were interviewed to explore the experience in implementing the information systems for maternal cases. Informants’ age range between 30 and 48 years-old. Four were midwives, 1 nurse, 1 general practitioner, 1 obstetrician, and 1 with a public health practitioner background.

The deductive analysis according to the framework with themes and sub-themes as seen in Table 2 below. The major challenges identified from policy analysis and the key informant interviews was the standard and monitoring, followed by quality of care, and communication. The policy analysis, confirmed by the interview, suggest that the availability of transportation, referral documentation and network infrastructure has not been significant challenge in conducting the maternal referral systems.

**Table 2.**
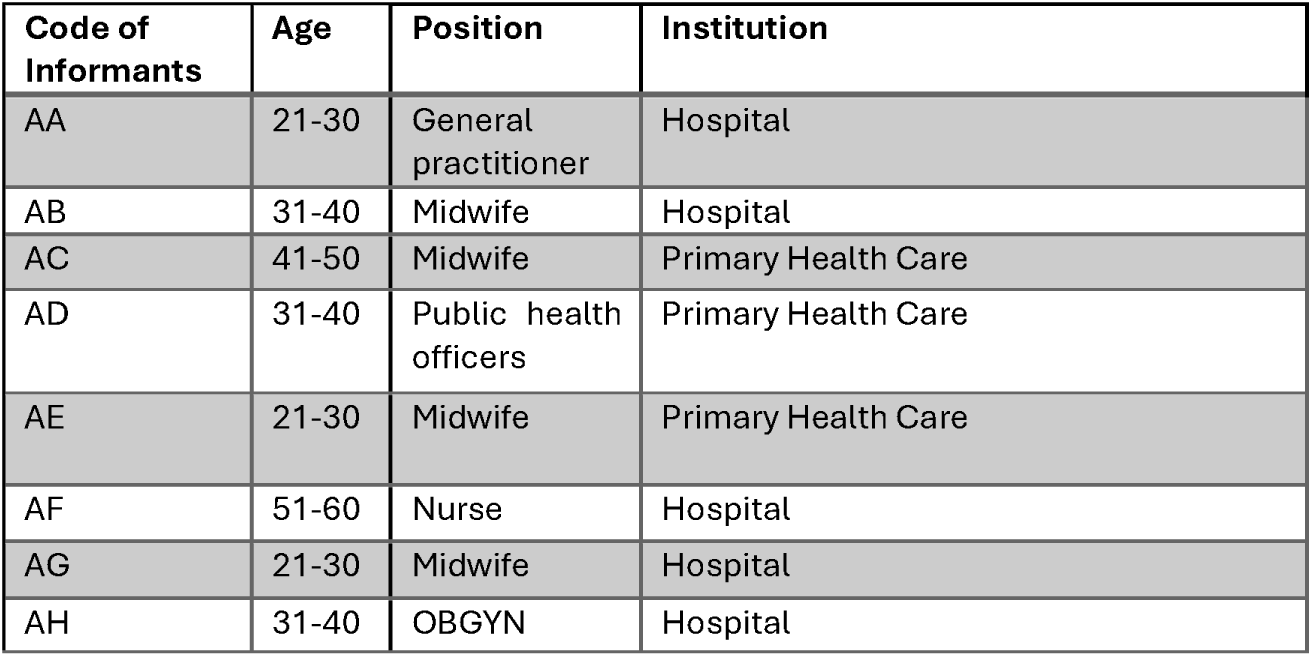
Study Informants.

**Table 3.** Themes and sub-themes of barriers in maternal and neonatal referral systems.

| Themes | Sub-themes |
| --- | --- |
| **Standard and monitoring** | Lack of case definition |
|  | Lack of monitoring and evaluation |
|  | Lack of health facility definition / facility network mapping |
|  | Limited monitoring activities from the authorities |
| **Communication** | Incomplete data from the referrers |
|  | Prolonged response time |
|  | Absence of guidelines for communication |
| **Quality of care** | Adequate number of health workers |
|  | Inadequate training for the health workers |
|  | Adequate equipment for setting up the e-referral systems |
| **Transportation** | Ambulance availability |
|  | Poor transportation access in selected area |
| **Referral documentation** | Good practice of documentation |
|  | Integration to electronic systems to record maternal data (e-maternal cohort) |
| **Network infrastructure** | Good network infrastructure |
|  | Good communication infrastructure |

### 1. Standard and monitoring

The policy analysis found that the existing information systems, Sisrute, SPGDT and Sitegar, do not have maternal case classifications or health facility classifications appropriate for maternal emergency management, such as BEONC or CEONC. Sisrute has a module for maternal emergency cases, however it does not describe whether the cases were applicable for ante-, intra-, or postpartum. This is confirmed by one of the respondents that mentioned that the Sitegar or Sisrute application only collect information payment methods and does not consider information on the maternal emergency cases classification.

> “For the classification of patients, they are usually asked about the region. and insurance whether social health insurance or cash payment (out-of-pocket). That was all the information required” (AC)

On the health facility classification, the Sisrute and Sitegar has the information on bed and equipment availability, but not for the SPGDT as its main purpose was to treat emergencies in the community. The three information systems did not possess information whether the hospitals have CEONC capacity or not. It was implied by one of the informants who were unsure about whether private hospitals have been assessed for their capacity of providing emergency care for maternal and neonatal. Further, there was no standard protocol of maternal referral pathway that has considered health facility capacity in providing emergency care for maternal and neonatal.

> “…if it’s district hospital, it’s definitely (qualified for) CEONC but I don’t know about private hospitals.” (AG)

Regarding the monitoring and evaluation, Sisrute has the dashboard to monitor systems utilization by the healthcare providers, however, limited monitoring or evaluation activity was conducted. For Sitegar, since its introduction in 2019, only one monitoring activity was conducted by the district health authority. The informant confirmed that an evaluation for Sisrute implementation was done once These 2 national referral systems could have been the more priority at that time that makes the use of SITEGAR becomes less of a concern.

> “Since the beginning of Sitegar there has never been an evaluation, at that time there was an evaluation of Sisrute. I am the doctor in charge of referrals (SPGDT) I have never observed any evaluation on the Sitegar.” (AA)

### 2. Communication

All the three information systems, Sisrute, SPGDT, and Sitegar has outlined the communication flow for referrals. The SITEGAR, for example, requires an operator that will respond to the text messages from referrers, ensure the bed availability, and confirm it to the referrers. The SITEGAR operator mentioned that the maximum initial response time to a text message is 5 minutes; however, this has yet to be formalized in the written guideline. The operator is a nurse or midwife that has been trained on the referral systems. The communication standard for SITEGAR operators and hospitals is a response time of less than 5 minutes as the initial response to receiving a referral but it has not been written in the operational standard explanation. After the patient is referred, the receiving facility will send the follow-up message on the patient’s condition during admission. Two sub-themes related to communication emerged from the transcripts: incomplete data from the referrers and the prolonged response time. The observation yields the third theme: absence of guidelines for communication.

The staff in charge of the SITEGAR at the hospital reported that one of the causes of long response time was the incomplete data. The SITEGAR pathway has determined the format of text messages for referral, however, the referrers may not complete the entire form. Moreover, some hospitals may require more information, such as the video of the patient’s condition, which makes the referrers need more time to provide it.

> “Sometimes it takes a long time because SITEGAR, for example, the referring midwife’s puskemas send to SITEGAR and SITEGAR sends to us, so yes, it turns out that there is incomplete data, for example regarding blood pressure, well then we ask what the blood pressure is to SITEGAR and SITEGAR conveys another question to the referring midwife and that’s a pretty long gap.” (AA)

The informant also reported that there are challenges in understanding and classifying the cases was not only because of lacking of case definition, but also due to the absence of video feature to allow the health professional at the recipient facility get a better understanding on patients’ condition. They usually used messaging platform to share information on patients’ conditions. While this could have been seen as something normal, but there is actually a factor of confidential passion data being spread through the messaging platform.

> “Yes, when we were on the way, we chatted with the referring midwife, via WhatsApp, we asked her to send a video because sometimes the hospital requested it.” (AG)

The observation to the examples of exchanged messages between the referrer and operator showed that one of the referrals, a case of a mother with preeclampsia that is going to have labor, was replied after more than 12 hours. The corresponding staff mentioned that one of the causes was the slow response from the destination hospital.

> “If you say it takes a long time, it’s quite a long time for urgent patients, for example, if we need the NICU, we can’t help but hurry up, especially since the opening is bound to increase and the contraction are good” (AE)

Further, the observation also found that the format of text messages varied considerably. It was implied that the variation is due to the lack of formal guidelines, for both primary healthcare providers (senders) and the hospital (recipients).

### 3. Quality of care

The analysis showed that each information systems has outlined the staff (health or non-health) that are involved, such as the operators and health personnel. However, there is still unclear standards of the training that the staff should receive, except for SPGDT that has already stated the different types of training for both health and non-health staff, including training on CEONC. The interview confirmed that the availability of health workers, midwives, was deemed sufficient in the Puskesmas and hospitals, including the ones served as referral systems operator. However, the training for emergency obstetric care was lacking.

> “Of the 16 midwives who work here, only 1 midwife had participated in the BEONC team training, if (training on) obstetric emergency, not all …” (AE)

> “For obstetric emergency, I think less than half of us had such training, and for integrated emergency care, or CEONC training, around 6 midwives had” (AB)

In terms of equipment, there was adequate tools or hardware required for setting up the Sisrute, SPGDT, and Sitegar systems at the hospital, each was done and used separate systems.

> “Yes, it’s true, the facilities are very adequate, there is its own room, wifi, tablets, tv stand by for referrals, electric generators and there is a 24-hour shift midwife, yes for SPGDT this referral is separate from the service.” (AB)

### 4. Transportation

Both the content analysis and interview demonstrated that the transportation systems have been adequate for the maternal referral purposes. Both selected Puskesmas have 1 two-wheeled mobile clinic, 1 four-wheeled mobile clinic, and one ambulance. Both Puskesmas also had dedicated drivers who were willing to stand by for 24-hours. This is confirmed by the following quotes.

> “It’s fine for the availability of ambulances, because there are two that usually used to refer, the driver is also there. Just be on call at night” (AE)

> Yes, we have 3 drivers so the shift system is 24 hours so if needed it is ready, from the petrol and manpower.” (AD)

The second sub-theme was road infrastructures. The midwives and staff at the district health authorities confirmed that some of both rural and urban areas have their challenges in road infrastructure, albeit differently. While the rural areas have some road segment with poor quality, the urban areas are struggling with the traffic.

> “Yes, for example in the Jn and Jg areas, it is very far away, there are still many bad access roads.” (AG)

> “Yes, there are still bad roads, because many trucks usually pass the highway, so sometimes it is quite difficult for us to get to the hospital, especially if there is a traffic jam.” (AE)

### 5. Referral documentation

The analysis showed that, when using the information systems, the referral documentation will be automatically kept in the records. The interview also confirmed that the staff were confident that they had done the good practice of case documentation and there is no risk of data lost. All the data was kept in the server in Bogor Regency is well maintained and adequate.

> “We have never lost documentation because we wrote it down in one dedicated sheet on the emergency obstetric care” (AA)

The data collected has also already integrated with the electronic systems to record maternal data in Indonesia (e-cohort).

> “there is data on pregnant women giving birth, we have entered it into the e-cohort and we also backed it up with manual.” (AD)

### 6. Network infrastructure

Based on the policy analysis and interview excerpts, we conclude that the communication and network infrastructures were adequate. The telephone lines and communication infrastructure dedicated to the referral system in Bogor Regency were available, both in the hospitals and the Puskesmas. All Puskesmas in Bogor regency has internet connection and all midwives and ambulance drivers in the selected Puskesmas has a smartphone, thus access WhatsApp or text messaging application. In the hospital, the computer, phone, and the internet also adequate.

> “There are two dedicated computers (for the referral systems) and 119, 1 computer WhatsApp web … and there is a mobile phone ..” (AG)

> “For BEONC purpose, we have a dedicated mobile phone, sometimes I use my personal mobile phone for the referral systems too. there is also safe WiFi connection.” (AE)

Each Puskesmas and hospital also backed-up by generator to ensure the electricity and connectivity works for 24-hour.

> “It’s rare (lost of connectivity) because the WiFi connection has a dedicated line (for the information and referral systems) also divided so it’s not slow, and we also have a gene set if the lights suddenly go out.” (AD)

All in all, the aspects of network infrastructures were deemed adequate by the informants interviewed.

## DISCUSSION

This is the first study that compares the multiple policies and information systems in place about patient referral and takes a real-world implementation example in one of the districts in Indonesia. The analysis showed that enacting the standards of maternal case definition, maternal emergency capacity of health facilities, and trainings for health and non-health workers is imperative to support an effective maternal emergency referral systems. This qualitative study identified that the key real-world challenges of utilizing information systems for maternal cases were lack of case definition and facility network mapping, limited monitoring activity, the incomplete data between the senders and recipients during pre-referral communication, prolonged response time, and inadequate standardization of maternal emergency training for health workers. Also identified was that no major challenges in terms of availability of vehicle for the referral, good practice of documenting the referral, and adequate network infrastructure.

The lack of clear standardized definitions of maternal cases could be the key challenges in ensuring the optimal use of SITEGAR in reducing maternal morbidities and mortalities. Such challenges align with those occurred in other LMICs (25, 26). Previous studies revealed the importance of maternal emergency triage systems in improving maternal morbidity and reducing mortality (27, 28). Such systems grouped maternal cases into level of severity and urgency in 5 levels: resuscitation, emergency, urgent, less urgent, and not urgent. While these studies applied the case categorization in the hospital-level, it can be beneficial to have this categorization applied in the case definition within the district-wide referral systems to allow a better identification of the capable health facility to receive the patients.

Furthermore, our study also identified the importance of identify, map, and develop a network of health facilities in an area to allow better and more prompt response in the referral systems. This, the mapping of network, had actually been exercised during the pilot of SijariEmas, however, it was not maintained after the pilot stages (14). Studies have recommended the applications of geographic information systems to improve healthcare accessibility, including for maternal and neonatal referral pathways (29–31). More recent one also highlights the importance of optimizing the machine learning in developing referral systems from primary healthcare facilities to hospitals (32). Such efforts could be developed further in strengthening maternal and neonatal referral pathways.

Our findings contradict the evidence from past studies in many LMICs that transportation and connectivity has been the key challenges in executing maternal referral pathways (26, 33, 34). This could be due to the study settings in urban areas, while majority of studies in LMICs were in rural areas. It implied that urban cities in Indonesia could have better opportunities in developing the referral of maternal and neonatal emergency cases and such efforts should be widespread in Indonesian cities with adequate network and road infrastructures.

While our study contradicts other LMICs-based studies that the number of health workers is limited, it agrees that the training for health workers is lacking (33, 35, 36). It is critical to introduce a comprehensive, not siloed, training for the health workers. In particular for the maternal referral pathways, the training should not only comprise of clinical skills, but also managerial and digital-related skills (37).

This study has several limitations. First, the study was focused on the health systems challenges of using the electronic referral systems for maternal and neonatal care, but not from the patient perspectives (i.e. environment, knowledge about referral, maternal health status, culture, and poverty). Concerning the evidence in other LMICs that individual and family factors among the key barriers of mothers referred to hospitals, future studies are warranted to discuss those topics. Second, the number of interviewed individuals is limited in this study. However, we used the maximum variation strategy, and these respondents are those considered most knowledgeable about implementation of SITEGAR for maternal and neonatal cases. We also conducted observation to explore the real-world challenge in executing the SITEGAR among the frontliners. Third, the study results were analyzed using the specific framework on the use of electronic referral systems for maternal cases, with limited emphasis on the human, organization, and technology fit (HOT-FIT) framework (38). We, however, used the widely-known framework that is tailored to the maternal referral cases so that it can put more emphasis on the issues related to maternal referral pathways.

This study contributes to the growing body of literature on digital health interventions in LMICs by highlighting both the policy content and the real-world challenges of implementing electronic referral systems for maternal care. Previous frameworks, such as the WHO’s Classification of Digital Health Interventions and the *HOT-FIT* framework, emphasize the importance of technological, organizational, and human factors in the successful adoption of digital health solutions. Our findings align with these models and propose the importance of policy factors, especially in the gaps in case definition, standardization, and training. These insights provide a foundation for designing more sustainable and scalable digital referral systems in LMICs.

## CONCLUSION AND RECOMMENDATION

The use of information systems for effective maternal healthcare referral remains a challenge in Indonesia, and the challenges were not only about the real-world implementation but also the content of the policy. To improve the effectiveness of referral systems, it is critical to define the clear maternal emergency case definition and classification, map the maternal emergency capacity of primary and referral-level healthcare facilities, set the basic competences that should be achieved from the Key barriers hindering the seamless use of maternal referral system, including the lack of standards and monitoring, limited formalized guidelines on the case definition and health facility network mapping, absence of communication standards, and limited and siloed training for the health workers. To improve the usefulness of SITEGAR in improving maternal morbidities and reducing mortalities, we recommend that the district or provincial health authorities in Indonesia to develop local referral systems by determining the category of maternal cases according to severity and urgency, map the existing healthcare facilities to fit (38) the capacity in maternal case management, establish formal guidelines on the communication procedure, and provide an integrated training that is not only focusing on clinical skills, but also the managerial and digital and technological. Health authorities should conduct more routine monitoring and used the information obtained from the monitoring process to improve the program implementation.

## Data Availability

Data are available upon request to the author.

## ACKNOWLEDGMENTS

We would like to thank all informants who participated and shared their thoughts and experiences for the study. This study is supported by Final Project Recognition Grant Universitas Gadjah Mada Number 5075/UN1.P.II/Dit-Lit/PT.01.01/2023.

## AUTHOR CONTRIBUTIONS

SA, LT, and LP designed the study. SA analyzed the data. SA and LP draft and wrote the manuscript. LT provides feedback to shape and articulate the key messages. The authors read and approved the final manuscript.

SA= Siti Angelina, LT=Laksono Trisnantoro, LP=Likke Putri.

